# Hospital and Environmental Transmission of XDR *Salmonella* Isangi Revealed by Genomic Surveillance in Malawi and South Africa

**DOI:** 10.64898/2026.03.11.26348031

**Authors:** Peter I. Johnston, Allan Zuza, Oliver Pearse, Erin M. Vasicek, Belson Kutambe, Happy Banda, Jonathan Rigby, Kenneth Chizani, Catherine Wilson, Priyanka D. Patel, Catherine Anscombe, Nathan J. Raabe, Lora Lee Pless, Kady D. Waggle, Lee H. Harrison, Shareef Abrahams, Juno Thomas, Phuti Sekwadi, Samantha Lissauer, Kondwani Kawaza, Anthony M. Smith, Jay C. D. Hinton, John S. Gunn, Melita A. Gordon, Nicholas Feasey, Philip M. Ashton

## Abstract

**Background:** *Salmonella* Isangi is an under-characterised serovar repeatedly associated with antimicrobial resistant hospital infections. Outbreaks of extensively drug-resistant (XDR) *Salmonella* Isangi occurred in close succession within hospitals in Malawi and South Africa, prompting us to characterise the serovar using epidemiologic, phenotypic, and genomic perspectives.

**Methods:** In Malawi, we integrated hospital blood culture surveillance with environmental sampling from neonatal wards and urban waterways. In South Africa, we analysed isolates from five hospitals involved in a regional outbreak. We used whole genome sequencing (Illumina and MinION) to characterise AMR genes and plasmids, assessed biofilm formation, disinfectant susceptibility, in *vivo* virulence, and analysed all publicly available *Salmonella* Isangi genomes.

**Findings:** 224 / 345 (65%) of genomes in the global collection belonged to *Salmonella* Isangi sequence type (ST) 335. Of these, 221 (99%) originated from Malawi and South Africa, including the isolates recovered from both outbreaks. 199 (89%) ST335 genomes carried determinants of resistance to fluoroquinolones and third-generation cephalosporins, consistent with an XDR profile.

In Malawi, a single ST335 clade caused the outbreak and was simultaneously present in both the hospital environment and nearby rivers. Inter-hospital transmission of a separate ST335 clade sustained the outbreak in South Africa. Closely related Malawian and South African isolates carried distinct plasmids encoding similar resistance determinants; evidence from our study and public databases suggests gene transfer via a cointegrate intermediate. Five non-outbreak South African ST335 isolates harboured additional carbapenem and macrolide resistance genes. Phenotypically, *Salmonella* Isangi ST335 resembled *Salmonella* Typhimurium in biofilm formation and disinfectant tolerance but was less virulent in mice.

**Interpretation:** *Salmonella* Isangi ST335 combines a locally untreatable XDR profile with nosocomial transmission and environmental persistence, suggesting a high potential for future outbreaks. A distinct and potentially greater threat lies in the horizontal spread of its resistance determinants to *Salmonella* Typhimurium and *Salmonella* Enteritidis, the two dominant invasive serovars in the region. Strengthened surveillance, integrating phenotypic testing with targeted genomics, is urgently needed. Its absence in Malawi, in contrast to South Africa, underscores inequities in preparedness for emerging AMR threats.

**Funding:** This work was supported by the Wellcome Trust through the Core Grant (206545/Z/17/Z) and the COVID-19 Sequencing Grant (220757/Z/20/Z) awarded to MLW. Additional support was provided by the Global Health Research Professorship to Melita Gordon from the UK National Institute for Health and Care Research (NIHR) (NIHR300039). Peter Johnston is funded by the Liverpool Clinical PhD Programme for Health Priorities in the Global South, supported by the Wellcome Trust (223502/Z/21/Z). For open access, the author has applied a CC BY public copyright license to any author-accepted manuscript version arising from this submission.

Whole-genome sequencing of *Salmonella* isolates from South Africa was made possible by support from the SEQAFRICA project which is funded by the Department of Health and Social Care’s Fleming Fund using UK aid. The views expressed in this publication are those of the authors and not necessarily those of the UK Department of Health and Social Care or its Management Agent, Mott MacDonald.

Analyses in this study were supported in part through use of software and workflows developed under National Institute of Allergy and Infectious Diseases, National Institutes of Health (NIH) grant R21AI178369. The NIH had no role in study design, data collection, analysis/interpretation, or publication decisions.

**Research in Context:** *Evidence before this study:* *Salmonella* Isangi is a recurrent cause of antimicrobial resistant hospital outbreaks. We searched PubMed for *Salmonella* Isangi and related synonyms (to February 23rd, 2026) and identified 39 articles. No prior studies have examined transmission routes or provided phenotypic characterisation beyond antimicrobial resistance testing. Outbreaks have been reported from five hospitals on three continents, as well as a foodborne outbreak in China. Two major sequence types (STs) consistently appeared: ST335 and ST216. ST216 was widely geographically distributed and recovered from a variety of animal, meat, and environmental sources. ST335 was primarily associated with human clinical cases.

*Added value of this study:* This investigation was motivated by an outbreak of extensively drug-resistant (XDR) *Salmonella* Isangi at a hospital in Malawi. The Malawian outbreak occurred shortly before a multi-centre nosocomial outbreak in South Africa, and we provide insights from both in our analysis. We combined local epidemiology, phenotypic analyses, and global genomic characterisation to deliver a comprehensive description of the serovar. Both outbreaks were caused by ST335, which is the dominant sequence type in South Africa and Malawi, but by distinguishable clades in each country. In Malawi, genetically indistinguishable isolates were simultaneously circulating among patients, the hospital environment, and rivers throughout Blantyre City. Transfer of patients between hospitals is likely to have sustained the outbreak in South Africa. Recombination through a cointegrate intermediate may explain why the same resistance determinants are carried on distinct plasmid backbones within the Malawian and South African ST335 clades. We identify five ST335 isolates in South Africa that were not related to either outbreak and which harbour carbapenem and macrolide resistance genes in addition to an XDR genotype.

*Implications of all available evidence:* XDR *Salmonella* Isangi ST335 is a major threat in Malawi because effective therapy requires antibiotics that are seldom accessible in routine care. The ability of ST335 to transmit in hospitals and to persist in the environment may increase the risk of future outbreaks. *Salmonella* Isangi readily acquires and maintains antimicrobial resistance determinants through diverse plasmid backbones and recombination, raising concern for transfer to locally prevalent invasive *Salmonella* serovars. National genomic surveillance of the kind that exists in South Africa is essential to track and contain further resistance emergence, but such surveillance does not exist in Malawi. There is an urgent need to expand genomic surveillance in low-income countries if the threat posed by *Salmonella* Isangi and other pathogens that drive antimicrobial resistance is to be recognised early and effectively contained.

## INTRODUCTION

Invasive non-typhoidal *Salmonella* (iNTS) infections are a major cause of illness and death across Africa.^1^ *Salmonella enterica* subspecies *enterica* serovars Typhimurium and Enteritidis dominate this clinical landscape, commonly showing multidrug resistance (MDR).^2^ Extensively drug-resistant (XDR) strains, characterised by MDR plus resistance to fluoroquinolones and third-generation cephalosporins, remain less common.^3,4^ In many settings where iNTS disease is endemic, XDR strains will be untreatable using routinely available antibiotics.^5^ Here, we characterised XDR *Salmonella enterica* subspecies *enterica* serovar Isangi (*Salmonella* Isangi) in the context of two outbreaks in Malawi and South Africa.

In 2020, clinicians at Queen Elizabeth Central Hospital (QECH), Malawi, observed a concerning rise in bloodstream infections caused by a highly resistant *Salmonella* species. The infections occurred mainly in infants and in neonates who had never left the hospital, signalling nosocomial transmission. In South Africa, public health officials at the National Institute for Communicable Diseases (NICD) reported a healthcare-associated outbreak of *Salmonella* Isangi from March to September 2022. The South African isolates produced extended-spectrum β-lactamases (ESBL) and were resistant to ciprofloxacin. Frequent patient transfers and asymptomatic carriers were suspected to have enabled inter-hospital spread.^6^ In 2023 national surveillance data, *Salmonella* Isangi ranked third among pathogenic *Salmonella enterica* serovars in South Africa, underscoring its growing clinical relevance.^7,8^

*Salmonella* Isangi has caused hospital outbreaks since at least 1969, frequently involving strains resistant to multiple antibiotics and producing ESBL.^9–14^ However, the associated transmission pathways, environmental reservoirs, and mechanisms of resistance remain poorly understood. Here, we use whole genome sequencing, detailed phenotyping, and comprehensive environmental and clinical surveillance to profile the serovar in the context of outbreaks in Malawi and South Africa. We provide a global genomic context for these outbreaks, uncovering critical gaps in surveillance and control measures for this under-recognised drug-resistant pathogen.

## METHODS

### Operational Context of Outbreaks

We identified the Malawian outbreak through blood and cerebrospinal fluid (CSF) culture surveillance, which the Malawi-Liverpool-Wellcome Programme (MLW) has conducted at QECH since 1998. We identified *Salmonella* Isangi isolates from blood and CSF cultures collected between 28 May 2018 and 9 February 2023. Concurrently, we recovered isolates from environmental sampling as part of two separate research studies: one investigating transmission of resistant *Enterobacteriaceae* within the neonatal unit (July 2018–March 2020), and another surveying *Salmonella* contamination in river water and wastewater across Blantyre (June 2018–December 2020).^15^

In South Africa, the National Institute for Communicable Diseases (NICD) collected isolates from an outbreak occurring between 17 March and 19 September 2022 across four hospitals in Eastern Cape Province and one in neighbouring KwaZulu-Natal. NICD defined cases as positive cultures obtained ≥48 hours post-admission or following inter-hospital transfers.

### Whole Genome Sequencing

We performed short-read sequencing of Malawian clinical and neonatal ward environmental isolates using the Illumina MiSeq platform at MLW. We sequenced selected isolates using Oxford Nanopore MinION technology (rapid barcoding kit SQK-RBK114.24, guppy basecalling v6.5.7).^16^ Isolates collected from river water were sequenced at the Wellcome Sanger Institute as described previously.^15^ As part of surveillance efforts, the NICD, South Africa (through the NICD Sequencing Core Facility), sequenced human isolates using the Illumina NextSeq platform.

We provide DNA extraction and sequencing protocols in the Supplementary Appendix.

### Publicly Available Genomes

We retrieved additional *Salmonella* Isangi genomes from EnteroBase and NCBI GenBank. On 2 April 2024 we queried EnteroBase with “Serovar = Isangi OR ST in (216, 335, 1994, 7036, 2261, 3390, 369, 5028)”, obtaining 281 genomes; 274 of these had available reads in the NCBI Sequence Read Archive, which we downloaded using Entrez Direct (v22.4).^17^

### Bioinformatic Analyses

#### Genome Assembly and Analysis

We trimmed raw Illumina reads with BBDuk (v38.9)^18^ and assembled them using SPAdes (v4.0.0)^19^. We excluded genomes exceeding 5.6 Mbp or 300 contigs based on QUAST (v5.3.0)^20^ results. We identified serovar, Multilocus Sequence Type (MLST), antimicrobial resistance (AMR), and virulence genes using SISTR (v1.1.0)^21^, mlst (v2.22.0)^22^, and AMRFinderPlus with default parameters (v4.0.15)^23^, respectively. Pseudogenes were identified using Pseudofinder v1.1.0 using a protein sequence database from Bakta v5.1 (https://zenodo.org/records/14916843). To compare the number of AMR genes in *Salmonella* (non-Isangi), *Salmonella* Isangi (ST335), and *Salmonella* Isangi (non-ST335) we used the “AMR genotypes” from the NCBI Pathogens portal (https://www.ncbi.nlm.nih.gov/pathogens/).

#### Reference Genome and Phylogenetics

We constructed a high-quality reference genome from Illumina and MinION sequencing data of a Malawian bloodstream isolate using Flye (v2.9.5)^24^, Medaka (v2.0.1)^25^, and POLCA (v3.4.1)^26^. We mapped reads against the reference using bwa mem and called SNPs using GATK v3.8-1-0 in unified genotyper mode,^27^ filtered out probable recombination derived variants using Gubbins (v3.4)^28^, and built a maximum likelihood phylogeny with RAxML-NG (v1.2.2; GTR+Γ model, 1000 bootstrap replicates)^29^. We analysed population structure using RHierBAPS (v1.1.3)^30^.

#### Plasmid Analysis

We performed plasmid replicon typing on short read assemblies using mob_typer (v3.1.9)^31^. For genomes with both long and short reads available, we assembled and polished long reads with Flye (v2.9.5)^24^ and Medaka (v2.0.1)^25^, and further polished with short read data using POLCA (v3.4.1). We circularised plasmids with circulator and constructed sequence-similarity networks using pling^32^ from long-read sequencing data.

### Phenotypic Assays

#### Biofilm Formation

We compared biofilm formation among ten Malawian *Salmonella* Isangi isolates, *Salmonella* Typhimurium ATCC14028, and a clinical ST19 *Salmonella* Typhimurium isolate. We grew biofilms at 25°C, stained them with crystal violet, and quantified biomass at OD_570_. We visualized colony morphologies indicating cellulose and curli expression on YESCA and LB agar supplemented with Congo red, Coomassie brilliant blue, or calcofluor white, and used confocal microscopy with Syto9, calcofluor white, and anti-curli antibodies.

#### Mouse Infection Models

We assessed virulence and chronic carriage by infecting BALB/c (NRAMP1-negative) intra-gastrically, and quantified bacterial loads from organ homogenates post-infection.

#### Antimicrobial Susceptibility

We determined minimum inhibitory concentrations (MICs) and minimum bactericidal concentrations (MBCs) for hospital disinfectants (bleach, chlorine, chlorhexidine) using broth microdilution and agar spotting.

Detailed methods for the three phenotypic assays described above are provided in the Supplementary Materials.

#### Ethical Approval

Malawian studies were approved by the College of Medicine Research Ethics Committee (COMREC approvals P.10/18/2499 and P.07/20/3089) and sponsored by the Liverpool School of Tropical Medicine. Reuse of sub-cultured bacterial isolates from these studies has been additionally approved (COMREC reference P06/20/3071). Mouse care and housing was carried out in accordance with guidelines established by the Abigail Wexner Research Institute (AWRI) Institutional Animal Care and Use Committee (IACUC) with an approved protocol (AR18-00080). The research activity followed the practices outlined in the *Guide for the Care and Use of Laboratory Animals*.

For South African studies, ethical approval to perform surveillance activities and laboratory analysis on clinical isolates of *Salmonella* was obtained from the Human Research Ethics Committee of the University of the Witwatersrand, Johannesburg, South Africa (protocol reference numbers: M160667, M1809107, M210752, M230985).

## RESULTS

### Hospital Outbreak in Malawi

We identified 21 *Salmonella* Isangi isolates from normally sterile sites at QECH between 28 May 2018 and 9 February 2023. Two isolates were from CSF and 19 from blood cultures. All infections occurred in separate individuals: 15 (71%) were in infants younger than one year (the youngest aged one week), five (24%) in children aged one to 13 years, and one in a 26-year-old adult. Five of the neonatal cases occurred in infants hospitalised since birth (Table 1).

**Table 1:** Epidemiological information for 21 invasive Salmonella Isangi infections identified at Queen Elizabeth Central Hospital, Malawi.

| Age (months) | Type of invasive isolate | Ward location | Collection date | Clinical details (from culture forms) |
| --- | --- | --- | --- | --- |
| 7 | Blood culture | General paediatrics | May 2018 | Septic, dehydration |
| 0.2 | Blood culture | Paediatric intensive care | March 2019 | Gastroschisis repair |
| 32 | Blood culture | General paediatrics | May 2019 | Severe acute malnutrition |
| 7 | Blood culture | General paediatrics | June 2019 | Severe acute malnutrition, acute gastroenteritis, dehydration |
| 7 | Blood culture | General paediatrics | June 2019 | Severe acute malnutrition, fever, dermatosis |
| 0.3 | Blood culture | Neonatal unit | December 2019 | Preterm, neonatal sepsis, hypoglycaemic |
| 0.5 | CSF | Neonatal unit | January 2020 | Suspected meningitis |
| 0.5 | Blood culture | Neonatal unit | July 2020 | Preterm, respiratory distress syndrome, sepsis, on CPAP |
| 156 | Blood culture | Paediatric medical | March 2020 | Kaposi's sarcoma |
| 0.7 | Blood culture | Neonatal unit | March 2020 | Prematurity sepsis |
| 1.6 | Blood culture | Paediatric nursery | April 2020 | Neonatal sepsis |
| 312 | Blood culture | Admissions unit | September 2020 | No clinical details given |
| 4 | Blood culture | Paediatric nursery | December 2020 | Nephrotic syndrome finished doses of ceftriaxone and metronidazole |
| 7 | CSF | General paediatrics | February 2021 | Suspected ventriculitis |
| 7 | Blood culture | General paediatrics | February 2021 | Hydrocephalus, ventriculitis |
| 6 | Blood culture | General paediatrics | December 2021 | Sepsis, cardiac failure, severe acute malnutrition, pneumonia |
| 9 | Blood culture | Paediatric High dependency | April 2022 | none provided |
| 40 | Blood culture | Paediatric intensive care | May 2022 | "on ceftriaxone + metronidazole" 3 days now |
| 5 | Blood culture | Paediatric oncology | October 2022 | No clinical details given |
| 15 | Blood culture | Paediatric Special Care Ward | December 2022 | fever |
| 28 | Blood culture | Paediatric admissions unit | February 2023 | Cerebral palsy |

Between 11 February and 30 March 2020, we isolated 37 *Salmonella* Isangi samples from the neonatal ward environment. Sampling sites included sinks, cots, oxygen delivery devices, and other high-touch surfaces (see Supplementary Table 1).^33^ In addition, *Salmonella* Isangi was identified from the hands of three mothers caring for admitted neonates and from the stool of three neonates. All environmental isolates differed by three or fewer single nucleotide polymorphisms (SNPs), consistent with recent common ancestry and local transmission.

Four neonates with invasive infections had received care on the neonatal ward. Two invasive isolates were genetically indistinguishable from environmental isolates, and two differed by two SNPs.

Between 17 August and 20 October 2020, we isolated 16 *Salmonella* Isangi samples from river water around Blantyre.^15^ Five of these were genetically indistinguishable from ward environmental isolates; four of these were collected in the same month as the ward environmental isolates, and one was collected later, in September 2020. The remaining 11 river isolates differed by up to 13 SNPs. 11 river isolates clustered downstream of QECH, though four isolates originated from waterways distant from the hospital. Figure 1A shows the proportion of non-typhoidal *Salmonella* blood culture isolates from QECH between 2015 and 2023, and their phenotypic resistance profiles. Figure 1B(i–ii) show timelines of isolate collection and corresponding SNP distances; Figure 1C illustrates isolate relatedness via a minimum spanning tree; Figure 1D maps *Salmonella* Isangi isolate locations relative to QECH and non-Isangi *Salmonella* isolates within Blantyre City.

**Figure 1 A.**
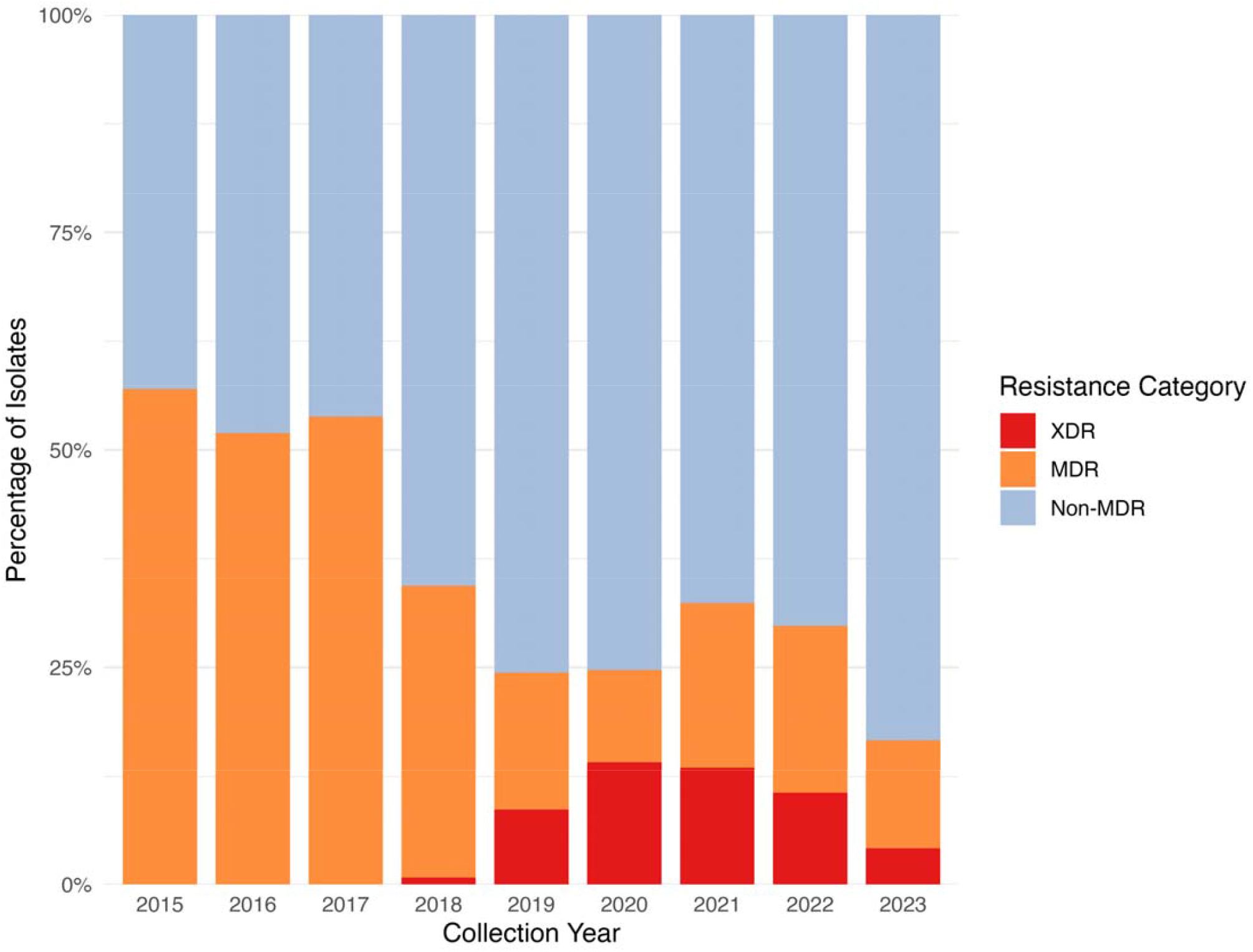
Phenotypic susceptibility of non-typhoidal Salmonella blood-culture isolates at Queen Elizabeth Central Hospital (QECH), Blantyre, Malawi, 2015-2023. Stacked bars show the yearly composition of isolates by susceptibility category: Non-MDR, MDR (resistant to ampicillin, chloramphenicol, and cotrimoxazole), and XDR (MDR plus resistance to cefpodoxime [used as a proxy for ceftriaxone] and a fluoroquinolone [ciprofloxacin or pefloxacin]).

**Figure 1 B.**
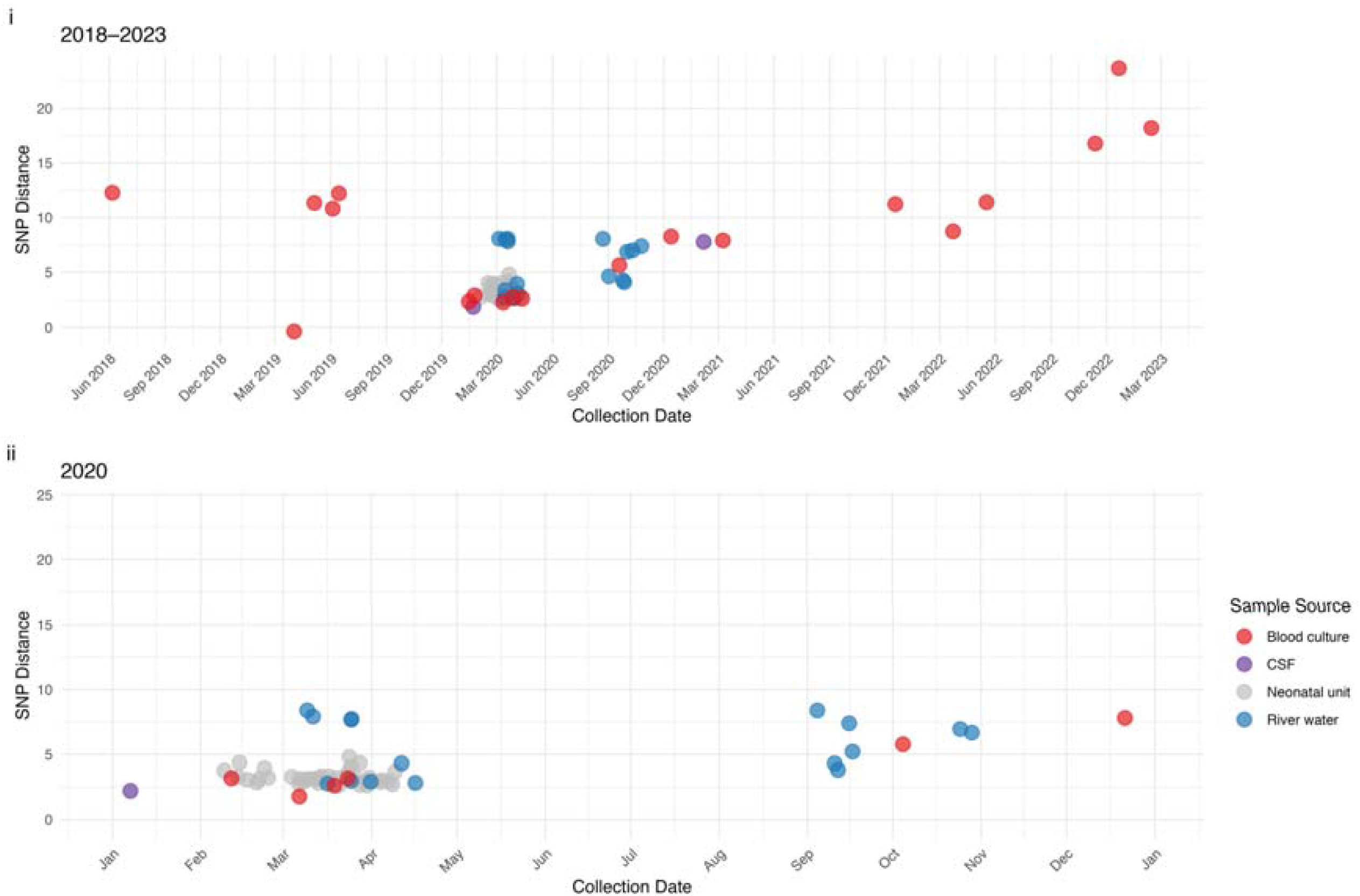
Temporal distribution of Salmonella Isangi isolates versus SNP distance to the reference blood culture isolate from the 2020 outbreak. (i) Isolates collected between May 2018 and February 2023, coloured by source type. (ii) Zoomed-in view of the 2020 outbreak period, highlighting the close genetic clustering of neonatal unit and river water isolates around the reference point. The dotted line indicates a 5-SNP threshold commonly used to infer close genomic relatedness.

**Figure 1 C.**
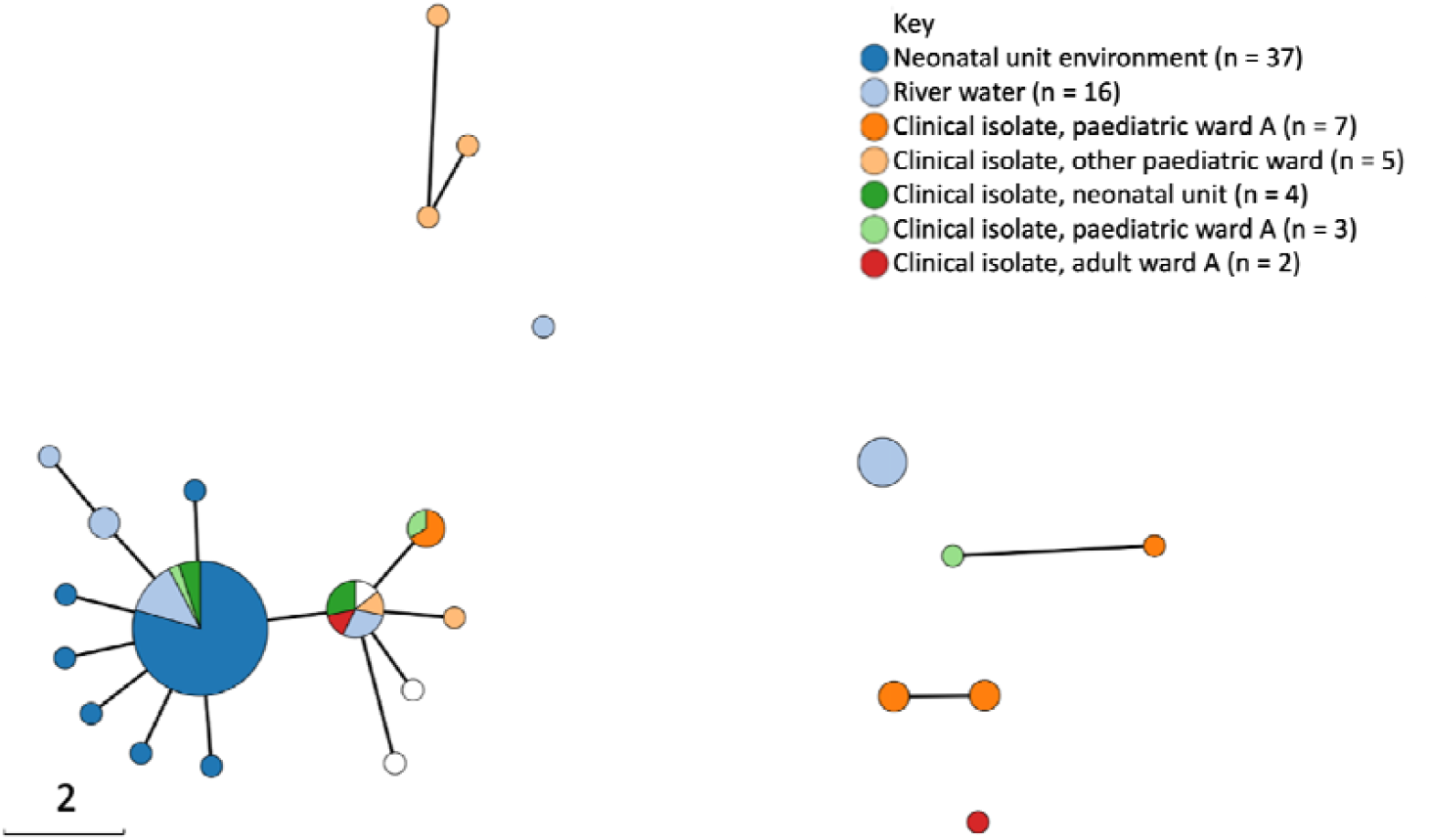
Minimum spanning tree, demonstrating single nucleotide polymorphism (SNP) distances between isolates from Malawi. Isolates within a circle are genetically indistinguishable. Edges between nodes represent SNP distances, as shown in the scale bar. Segments within circles are coloured by the origin of the constituent isolates: dark blue = neonatal unit environment (‘Env – CN’); light blue = river water; dark green = invasive isolates from babies on the neonatal unit (‘Human – CN’); orange = invasive isolates from children on the most frequently affected paediatric ward (‘Human – Moyo’); light green = isolates from babies on a nursery unit (‘Human – Nursery’); red = invasive isolates from adult patients (‘Human – Adult other’).

**Figure 1 D.**
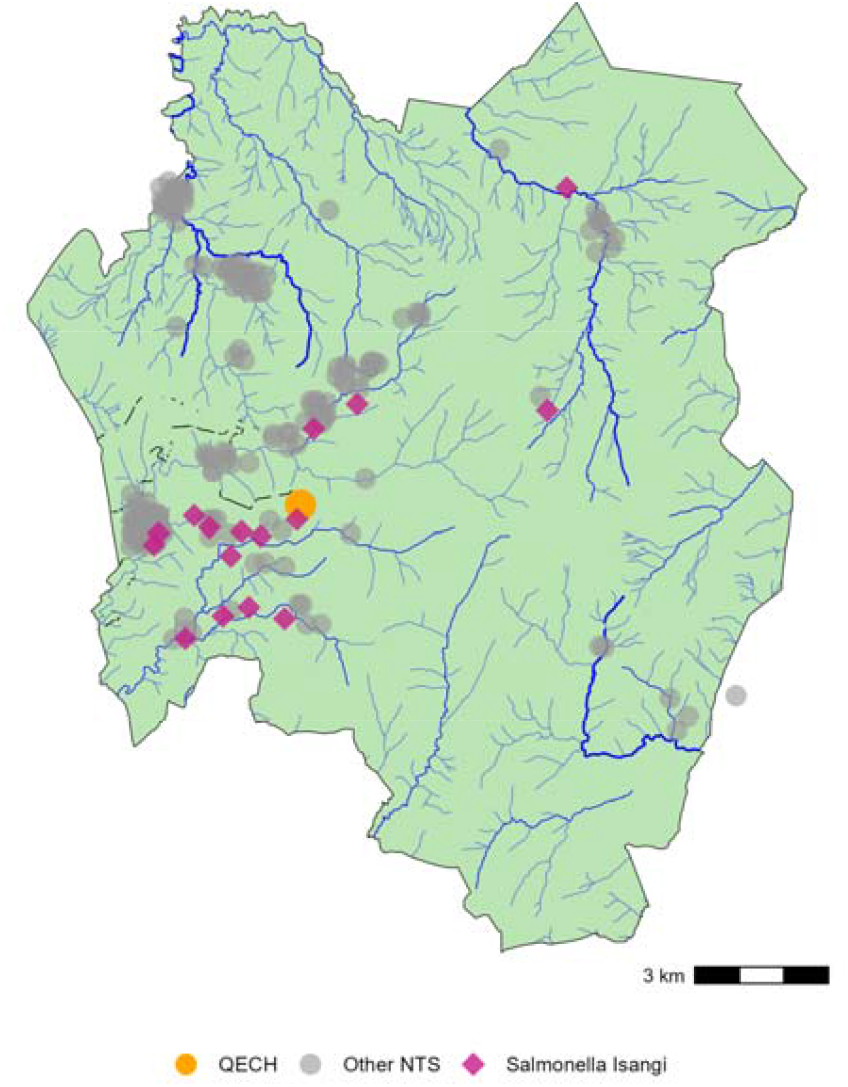
Map of Blantyre District showing locations of recovered Salmonella isolates in relation to Queen Elizabeth Central Hospital (QECH) and local waterways. Points represent the locations of Salmonella Isangi (purple diamonds) and other non-typhoidal Salmonella (NTS) isolates (grey circles).^15^ The location of QECH is marked with an orange circle. Waterways are shown in blue, with line width proportional to estimated flow rate.

**Figure 2.**
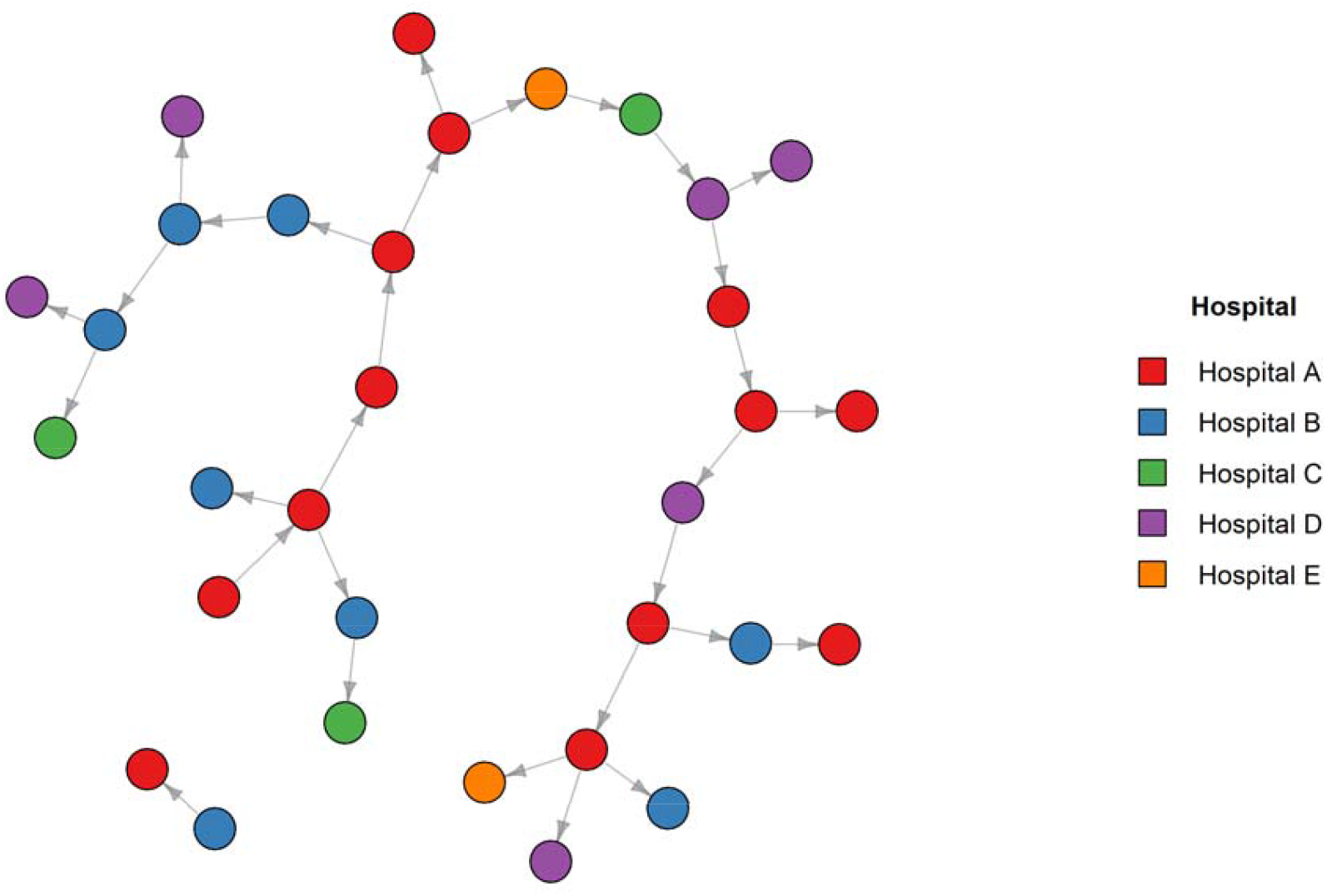
Reconstructed transmission tree for the Eastern Cape outbreak, created using outbreaker2 (v1.1.3). Nodes represent individual cases, colour-coded by hospital of origin: Hospital A (red), Hospital B (blue), Hospital C (green), Hospital D (purple), and Hospital E (orange). Directed edges indicate the most likely transmission pathways, with arrows pointing from the inferred source to the recipient. The network structure highlights multiple transmission chains, with cases distributed across hospitals, indicating inter-hospital transmission events.

### Hospital Outbreak in South Africa

Between 17 March and 19 September 2022, we collected 39 *Salmonella* Isangi isolates during a multi-facility healthcare-associated outbreak involving five hospitals in Eastern Cape Province, South Africa. Of these, 38 isolates (34 XDR) passed genomic quality control; four isolates originated from blood cultures, and the remainder came from stool or rectal swabs. Isolates formed a distinct phylogenetic clade interspersed with sporadic non-outbreak isolates from Eastern Cape and elsewhere in South Africa, with a maximum SNP distance of 108. Bayesian transmission reconstruction demonstrated repeated inter-hospital spread centred on one primary institution (Hospital A; *Error! Reference source not found*.).

### Population Structure and Global Phylogenomics

In total, we analysed 345 high-quality *Salmonella* Isangi genomes, of which 327 belonged to clonal complex 25 (CC25; Figure 3A). Within CC25 there was a phylogeographical split between ST335 (n=224), 99% of which were from Malawi or South Africa, and ST216 (n=102), which was detected in 12 countries across five continents. One additional genome was ST5028, a single-locus variant of ST216.

**Figure 3.**
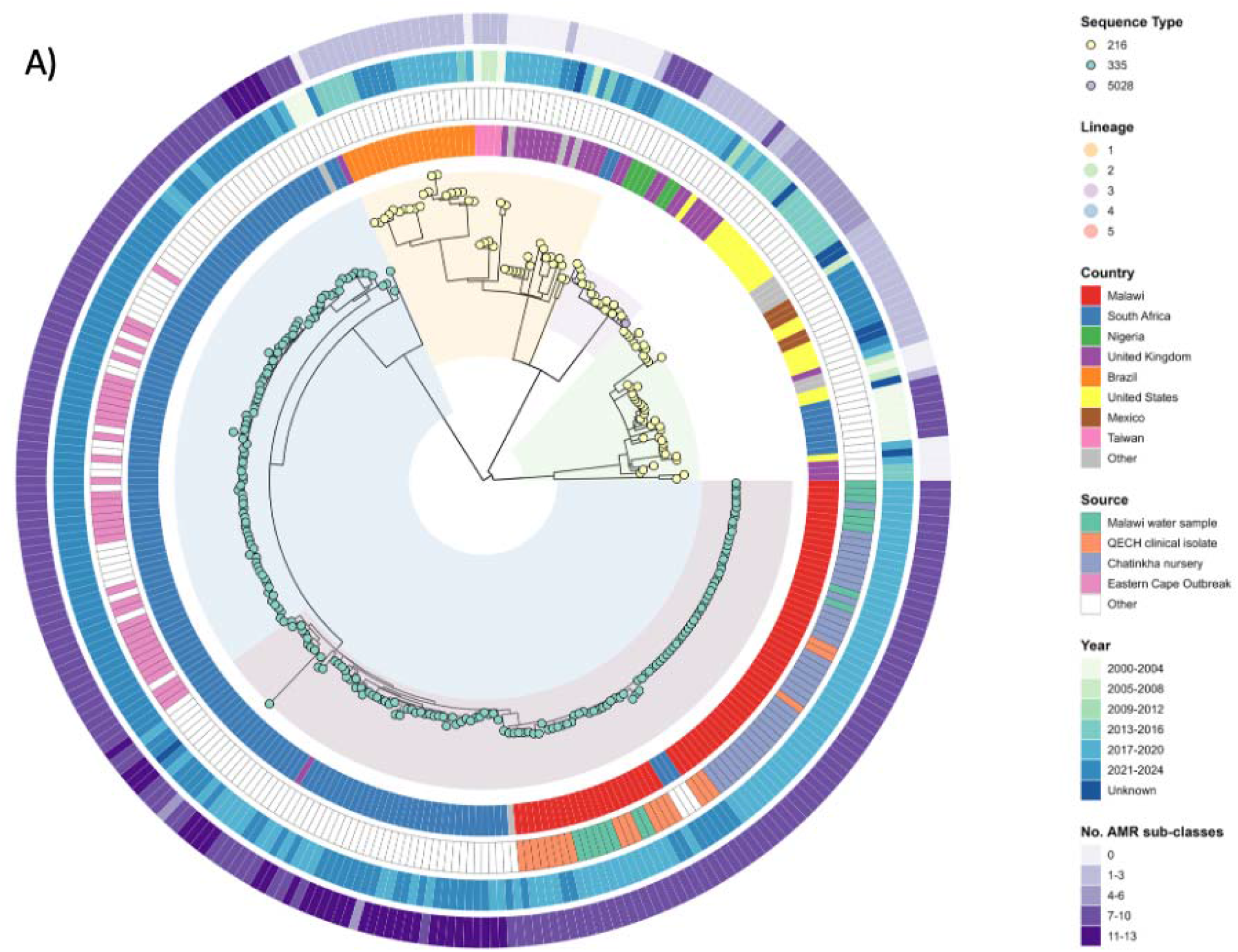
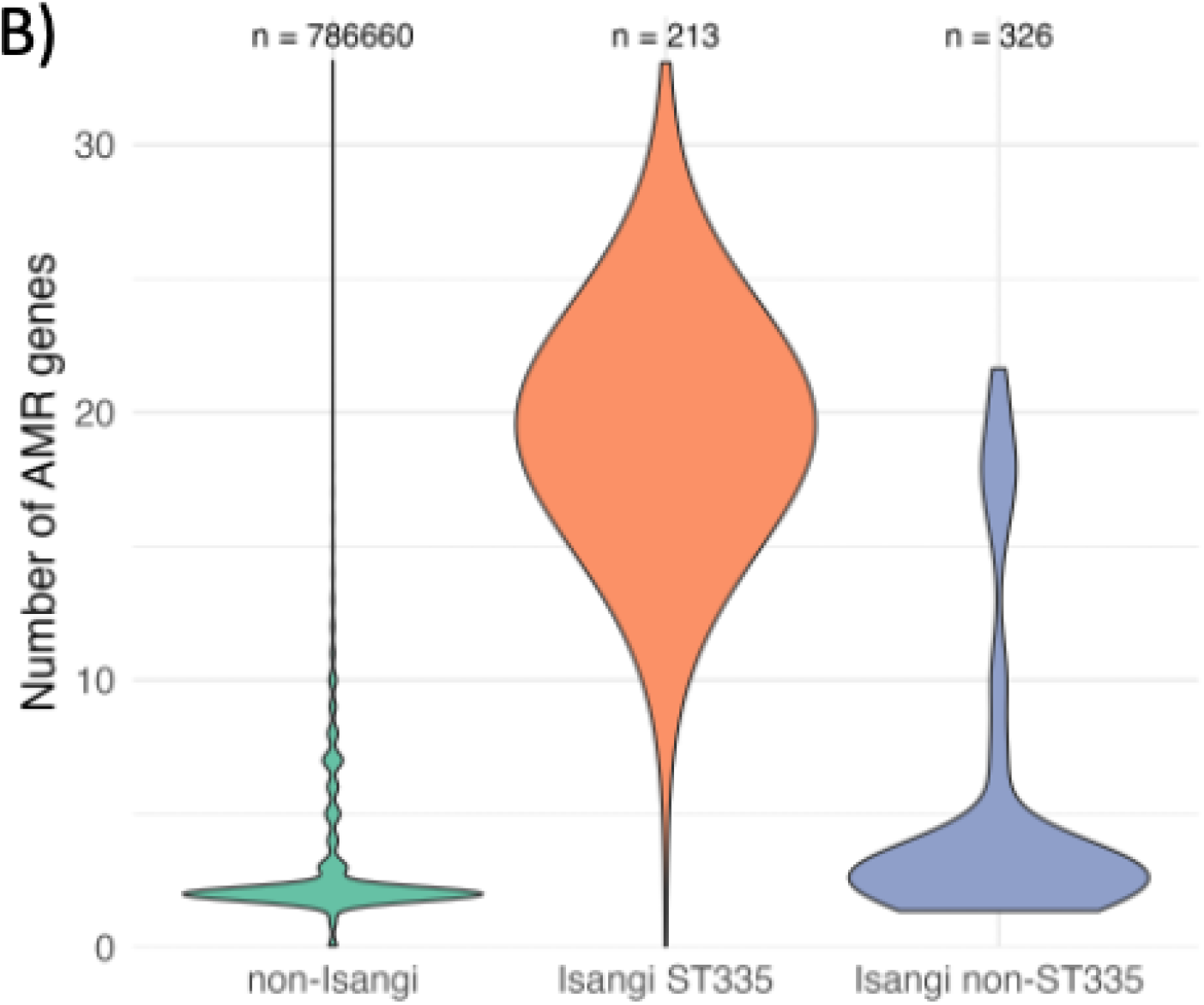
A) Comparison of the number of AMR genes in non-Isangi Salmonella, S. Isangi ST335, and S. Isangi that aren’t ST335 B) Maximum likelihood phylogenetic tree of Salmonella Isangi Clonal Complex 25 (CC25) isolates. Tree tips represent sequence type (ST), coloured blocks represent RHierBAPS lineage, while heatmaps (inner to outer rings) denote country of origin, year of collection and the number of antimicrobial resistance (AMR) subclasses. ST335 isolates are primarily from Southern Africa, while ST216 is more globally distributed. ST335 isolates show a higher frequency of antimicrobial resistance (AMR) sub-classes than ST216

**Figure 4.**
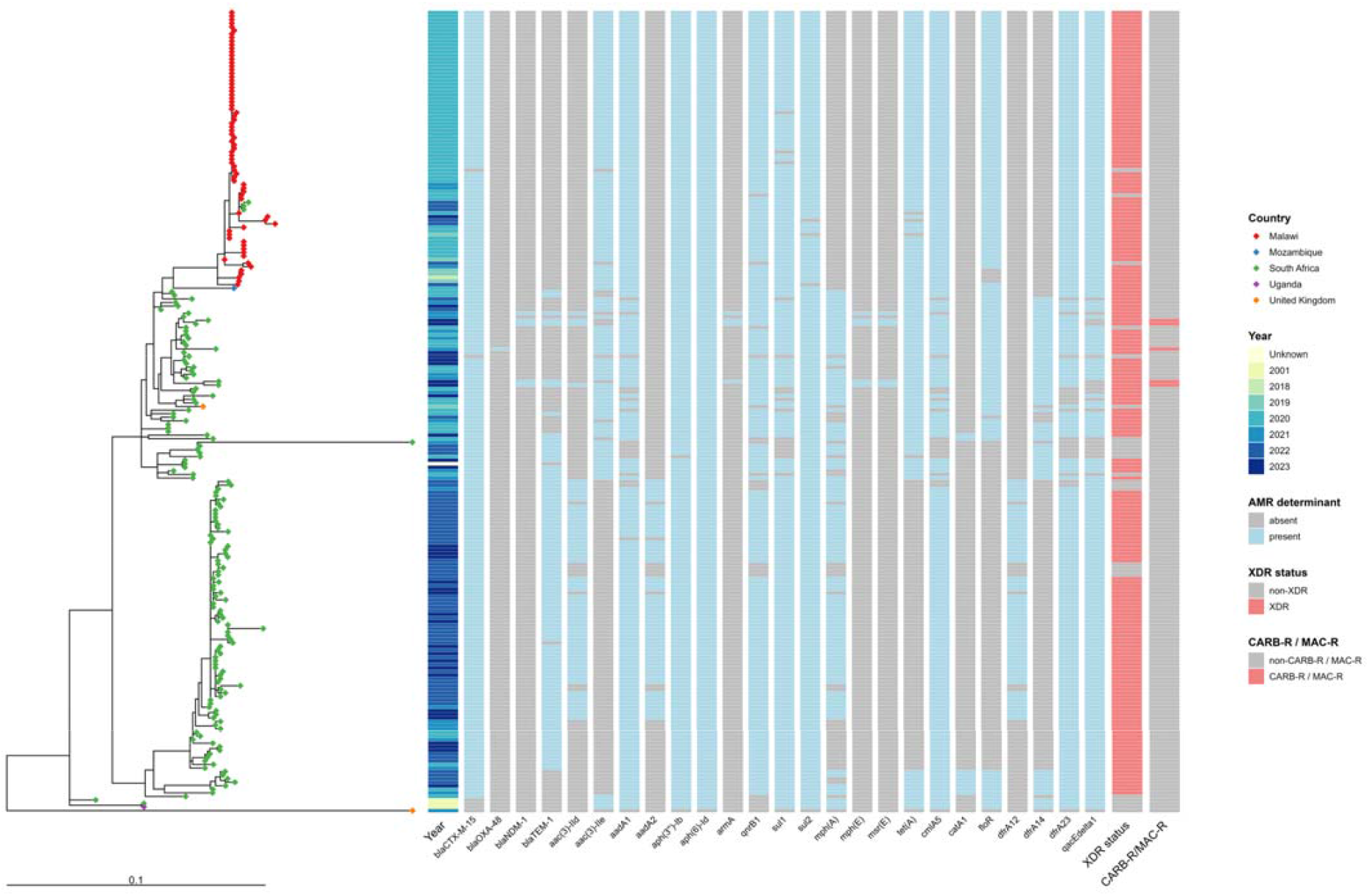
Midpoint-rooted maximum likelihood phylogenetic tree of Salmonella Isangi ST335 isolates. The tip colours represent the country of origin, with South African isolates in green and Malawian isolates in red. The Malawian isolates forming a distinct phylogenetic clade within the broader tree. The first column of the heatmap indicates the year of collection. The subsequent columns show the presence (light blue) or absence (grey) of key antimicrobial resistance (AMR) genes. The penultimate column defines genotypically predicted XDR status, based on resistance to amoxicillin, trimethoprim-sulfamethoxazole, chloramphenicol, fluoroquinolones, and third-generation cephalosporins. The final column indicates additional genotypically predicted resistance to carbapenems (CARB-R) and macrolides (MAC-R)

We defined five hierBAPS lineages within CC25. Lineages 1–3 were composed of ST216 genomes, while lineages 4 and 5 contained ST335 genomes. All 74 ST335 genomes from Malawi clustered within lineage 5. Of the 146 South African ST335 genomes, 55 (38%) belonged to lineage 5 and the remaining 91 (62%) to lineage 4. The Malawian lineage 5 isolates formed a distinct clade nested within a larger South African clade, consistent with a South African origin of the Malawian sub-clade. Of note, three South African isolates clustered within this Malawian sub-clade, suggesting regional transmission.

### Antimicrobial Resistance

We first compared the frequency of antimicrobial resistance genes across *Salmonella* populations. *Salmonella* Isangi ST335 carried a dramatically higher number of AMR genes than both non-ST335 Isangi and non-Isangi *Salmonella* (Figure 3B), identifying ST335 as an extreme outlier in resistance gene burden. Among ST335 isolates (n = 224), 199 (89%) possessed XDR genotypes, characterised by *bla*_*CTX-M-15*_ (ESBL determinant) and *qnrB*1 (fluoroquinolone resistance). A subset of 110 South African isolates encoded macrolide resistance (*mphA*), and five isolates from Gauteng Province (2021–2023) additionally carried *mphE, msrE*, and carbapenemases (*bla*_*NDM*-1_ or *bla*_*OXA*-48_). Fifteen ST335 isolates (7%) were MDR, while nine (4%) lacked certain classical MDR markers yet were resistant to ≥ 4 antibiotic classes. 205 (92%) of ST335 genomes carried the quaternary ammonium compound efflux transporter *qacEΔ1* gene. **Error! Reference source not found**. shows a phylogenetic tree of all ST335 isolates annotated with AMR gene determinants.

Among ST216 isolates (n = 102), seven (7%) were XDR (*bla*_*TEM-131*_ and *gyrA* D87N), all of which were isolated in South Africa, dating from 2001. A further eight (8%) ST216 genomes carried determinants conferring genotypic MDR; the remaining 75 (74%) of ST216 genomes harboured one or fewer resistance genes.

### Plasmid Characterisation

Plasmid replicon typing revealed that 150 of 155 (97%) South African CC25 genomes carried IncC plasmids, 27 (17%) carried IncHI2 plasmids, and 24 (16%) carried both replicons. All 74 Malawian genomes exclusively carried IncHI2 plasmids. Long-read sequencing of four representative isolates (SAL02348, CHF10J1, SAL01261, and SAL01259) confirmed distinct plasmid backbones: South African isolates carried AMR determinants within three genomic islands on IncC plasmids (1.8 kb, 6.9 kb, 5.2 kb), while the Malawian isolate carried four genomic islands (6.9 kb, 6.9 kb, 5.2 kb, 2.1 kb) on an IncHI2 plasmid (Figure 5). The South African isolates that were most closely phylogenetically related to the Malawian isolates carried both IncC and IncHI2 plasmids. Two islands were highly similar between plasmids, one showed partial conservation with rearrangements, and one was unique to the Malawian plasmids. We also identified a 444 kb hybrid IncC–IncHI2 plasmid (GenBank accession CP028197) in a *Salmonella* Concord isolate from food in Czechia (2018), which showed high similarity to the *Salmonella* Isangi AMR plasmids. This cointegrate plasmid had containment distances of <0.5 (i.e., at least 50% of the smaller plasmid sequence was contained within the larger plasmid for each comparison) and had four or less structural rearrangements (≤4 DCJ-Indel distance) compared to both the IncC (162 kb) and IncHI2 (204 kb) plasmids, suggesting that a plasmid co-integration event could have enabled spread of AMR genes in this case (**Error! Reference source not found**.).

**Figure 5.**
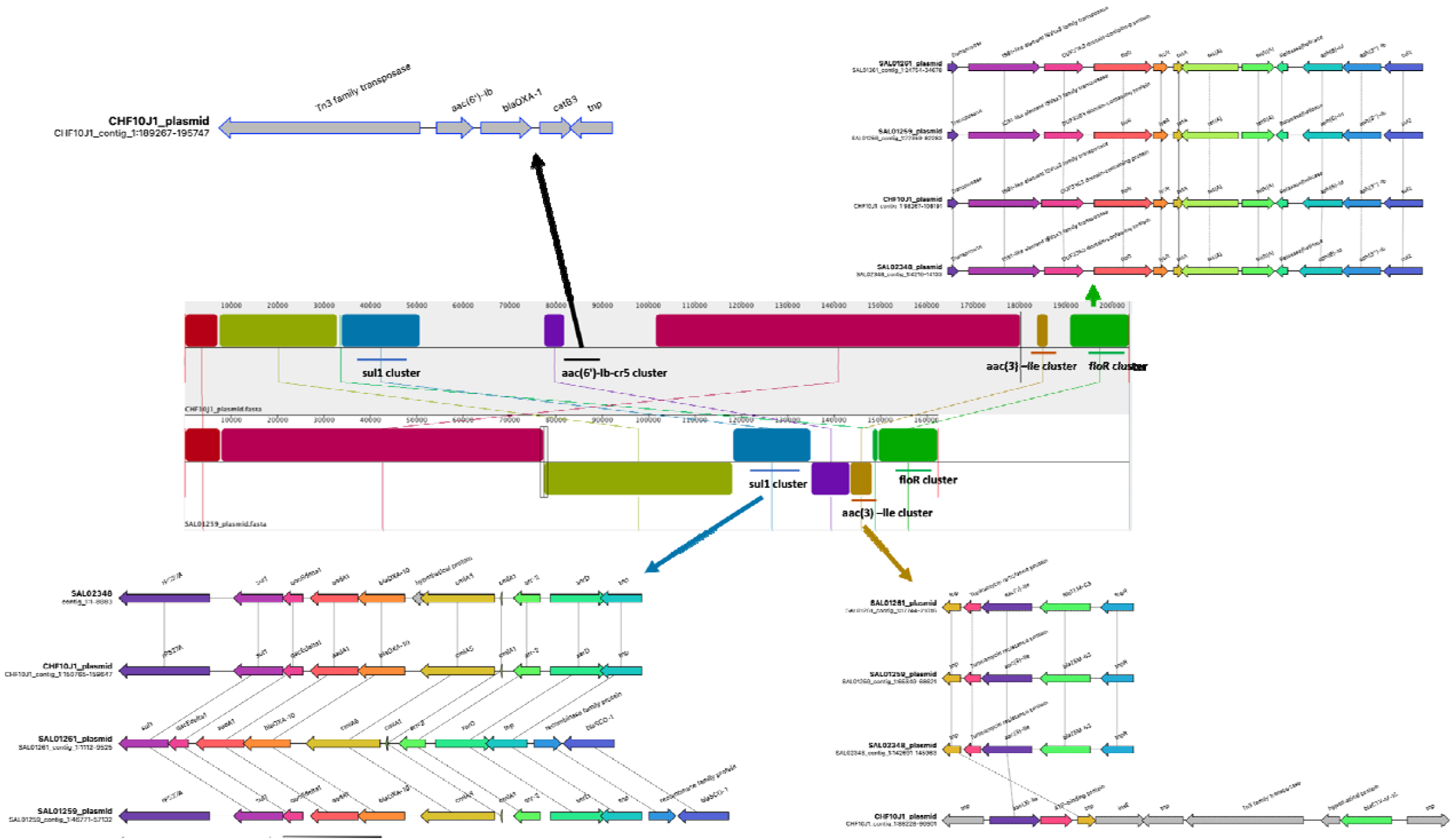
Comparative genomic analysis of AMR plasmids in South African and Malawian S. Isangi isolates. Central panel shows alignment of long-read-assembled IncHI2 (top) and IncC (bottom) plasmids from Malawian and South African isolates, respectively. Regions of similarity between the plasmid are shaded and linked by lines. Detailed gene structure is shown for the four AMR gene clusters, two of which are highly conserved (floR and sul1), one is somewhat conserved (aac(3)-IIe), and one which is unique to the Malawian isolate (aac(6’)-Ib-cr). Arrows denote genes, colour indicates same gene.

### Phenotypic Profiling

*Salmonella* Isangi ST335 isolates exhibited biofilm formation comparable to *Salmonella* Typhimurium in all tested conditions, though cellulose production was lower by biomass (Supplementary Figures 2 and 3) and on calcofluor white plates (data not shown). In mice, intragastric infections of BALB/c mice with *Salmonella* Isangi isolates resulted in no mortality, contrasting with *Salmonella* Typhimurium ATCC14028, which caused 50% mortality by day 14 (Supplementary Figure 4). Following this finding of reduced virulence, we investigated the presence of pseudogenes in the central anaerobic metabolism pathway, and found that *Salmonella* Isangi had an average of 27 disrupted genes in this pathway identified by Pseudofinder, similar to gastrointestinal *Salmonella* which had a range of 29 to 32 genes identified as disrupted by Pseudofinder in central anaerobic metabolism. Minimum inhibitory and bactericidal concentrations (MIC/MBC) for bleach and chlorine were identical across *Salmonella* isolates (Supplementary Table 2). *Salmonella* Isangi isolates demonstrated lower MICs to chlorhexidine relative to *Salmonella* Typhimurium.

## DISCUSSION

Here we characterised *Salmonella* Isangi in the context of two AMR-associated hospital outbreaks. ST335 was the predominant sequence type among global genomes, was localised to South Africa and Malawi, and caused both hospital outbreaks. 89% of ST335 genomes were XDR, and five non-outbreak isolates from South Africa had acquired additional determinants of resistance to macrolides and carbapenems. Plasmid recombination via a cointegrate intermediate could explain how Malawi and South Africa clades carried the same resistance determinants on distinct plasmid backbones. This could contribute to persistence within *Salmonella* Isangi and transfer of AMR genes into other invasive serovars.

In Malawi, the same XDR *Salmonella* Isangi ST335 clade was detected in paediatric invasive infections, the neonatal unit, and rivers downstream of the hospital, highlighting genomic continuity between clinical and environmental isolates. In South Africa, genomic and epidemiologic evidence indicated inter-hospital transmission of XDR ST335, highlighting the potential for recurrent outbreaks across multiple sites. Taken together, these findings indicate that the XDR profiles and detection of *Salmonella* Isangi in hospital and environmental samples could pose a public health concern.

This is the most comprehensive characterisation of *Salmonella* Isangi to date. We combine a large and geographically diverse dataset with high-resolution SNP-based genomic analysis, direct plasmid reconstruction, and integrated epidemiologic and phenotypic data from two prolonged outbreaks. This integrated approach allowed us to describe outbreak transmission and uncover plasmid recombination between IncC and IncHI2 backbones as a possible mechanism of AMR spread. Previous smaller genomic studies identified the endemicity of ST335 in South Africa and highlighted the near-ubiquitous presence of ESBL determinants such as *bla*_*CTX-M-15*,_ but did not include epidemiologic or phenotypic data or identify the plasmid-level processes driving resistance ^34–36^

Nosocomial *Salmonella* outbreaks are typically rare, foodborne, and transient.^37,38^ Our findings suggest that sustained hospital transmission could have been linked to environmental reservoirs. Similar situations have been reported previously; for example, fluoroquinolone-resistant *Salmonella* Schwarzengrund and Seftenberg in the United States and ESBL-producing *Salmonella* Livingstone in Tunisia. Such incidents remain uncommon enough to be noteworthy.^39–41^ Earlier reports relied on pulsed-field gel electrophoresis to show relatedness but did not identify genetic determinants. Our genomic approach enabled the integrated investigation of AMR determinants, genome degradation, and phylogenetic relatedness, suggesting the importance of AMR to sustain transmission in these settings.

There are striking similarities between the situation we report for XDR *Salmonella* Isangi and *Klebsiella pneumoniae*, which causes high levels of neonatal sepsis across Africa south of the Sahara. Both pathogens show extensive drug resistance, persistence in hospital environments, and infections in vulnerable populations.^42,43^ We identified up to four integrons carried on the same large plasmid in *Salmonella* Isangi, mirroring the plasmid-mediated integrons driving XDR *Klebsiella pneumoniae* outbreaks in African neonatal units. In this respect, *Salmonella* Isangi may exemplify how the same selective pressures that produced the so-called ESKAPE pathogens (*Enterococcus faecium, Staphylococcus aureus, Klebsiella pneumoniae, Acinetobacter baumannii, Pseudomonas aeruginosa*, and *Enterobacter* spp.) continue to drive the emergence of novel, hospital-adapted multidrug-resistant threats

Our data confirm plasmid-mediated AMR dissemination within *Salmonella* Isangi. Malawian and South African isolates had near-identical chromosomes and similar AMR genes, yet these genes were carried on different plasmid backbones. The earliest XDR *Salmonella* Isangi isolates from South Africa in our dataset carried an IncC plasmid, while more recent isolates had both IncC and IncHI2 plasmids. Malawian isolates exclusively carried AMR genes on IncHI2 plasmids. The detection of cointegrate IncC–IncHI2 plasmids in public databases raises the possibility that plasmid fusion events have driven AMR spread in *Salmonella* Isangi. Such events could increase the persistence and dissemination of resistance determinants and lead to transfer into other invasive serovars.

Critically, the mechanism through which *Salmonella* Isangi ST335 has acquired resistance could occur within other epidemiologically linked *Salmonella* serovars. *Salmonella* Typhimurium and *Salmonella* Enteritidis are the primary cause of invasive non-typhoidal *Salmonella* disease in the region, and loss of effective ceftriaxone therapy would likely increase the high death rate associated with these infections. Our findings emphasise the importance of identifying serovars such as *Salmonella* Isangi through genomic surveillance, so that threats posed by AMR spread from rarer serovars can be recognised and addressed.

While the relationship between resistance and virulence is complex, increased resistance has been associated with reduced virulence in certain bacterial pathogens.^44^ Our finding that *Salmonella* Isangi carries an unusually high number of resistance genes for a *Salmonella* isolate, yet showed very low virulence in a susceptible mouse model of systemic disease is consistent with a trade-off between resistance and virulence. However, this apparent attenuation *in vivo* contrasts with the ability of ST355 to cause bloodstream infections in patients in both Malawi and South Africa. One possible explanation for these divergent phenotypes is that *Salmonella* Isangi may be adapting toward human restriction. While we found no evidence of genome degradation to support this hypothesis alternative mechanisms could be responsible.^45^

Although *Salmonella* Isangi showed lower virulence than *Salmonella* Typhimurium in our susceptible mouse model of systemic disease, robust biofilm formation and tolerance to disinfectants raises the possibility of high environmental persistence and hospital spread. Rapid detection and strict infection control measures are important mitigations to put in place.

Genomic surveillance is critical for identifying and mitigating the spread of emerging AMR pathogens. In upper-middle-income settings like South Africa, national surveillance systems have identified *Salmonella* Isangi as the third most common non-typhoidal *Salmonella* serotype.^7^ However, in low-income settings like Malawi, detection has been primarily dependent on collaboration with research institutions, leaving the true burden of *Salmonella* Isangi and other resistant pathogens incompletely characterised.^46^ In the current climate of constrained global health financing, equitable surveillance will require pragmatic, tiered approaches that combine strengthened phenotypic testing at sentinel sites with targeted sequencing of high-risk isolates supported by regional reference laboratories.

Our findings were influenced by the substantial contribution of genomes from NICD, a clinically focused surveillance system in South Africa. This sampling structure likely over-represents invasive clinical isolates and under-represents community, environmental, and animal reservoirs, potentially biasing inferences toward hospital-associated transmission. In addition, genomic surveillance capacity differs markedly between South Africa and Malawi, meaning that cross-border comparisons may reflect differences in surveillance intensity rather than true differences in burden or lineage distribution. Finally, although whole-genome sequencing provides high resolution for assessing relatedness, genomic similarity alone cannot establish directionality of transmission without detailed epidemiological data. Future studies incorporating systematic sampling from community, environmental, and animal contexts across multiple settings would strengthen inference regarding the ecological niche and transmission dynamics of *Salmonella* Isangi.

The ST335 sequence type of *Salmonella* Isangi emerges intermittently in hospital settings where it can contaminate the environment and cause clinical disease. Although not dominant across the region, continued antibiotic selection pressure and surveillance gaps may facilitate further expansion. Our study highlights that serovars outside the usual invasive NTS paradigm can acquire and maintain extensive drug resistance and transition from episodic detection to sustained transmission. These findings reinforce the need for equitable genomic surveillance and strengthened infection control to identify and contain emerging AMR threats before they become more widely established.

## Data sharing

All the genomes sequenced as part of this study are available from European Nucleotide Archive under the project accession ERP189265.

## Data Availability

All the genomes sequenced as part of this study are available from European Nucleotide Archive under the project accession ERP189265, a full list of accessions is available in Supplementary Table 1.

## Acknowledgements

We thank all participants of the GERMS-SA Laboratory Surveillance Network, South Africa for submission of clinical isolates of *Salmonella* species to the National Institute for Communicable Diseases, South Africa. This work is dedicated to the memory of Elizabeth Ashton.

