## Supplementary Appendix for "Hospital and Environmental Transmission of XDR *Salmonella* Isangi Revealed by Genomic Surveillance in Malawi and South Africa"

**Supplementary Methods**

**Microbiologic methods for recovering salmonellae in Malawi**

**Microbiology**

Two sample types were collected for microbiology analysis; rectal swabs and stool samples, and ward surface swabs. Rectal swabs were directly plated on ESBL chromogenic agar (CHROMagar™ ESB, https://www.chromagar.com/en/) and incubated at 37 ℃ for 18 to 24 hours. Ward surface swabs were first incubated in 5ml of buffered peptone water (BPW) at 37 ℃ for 18 – 24 hours, after enrichment in BPW, 10μl of this sample was plated on ESBL chromogenic agar at 37 ℃ for 18 – 24 hours. Colonies from the ESBL chromogenic agar suspected to be *Salmonella* spp. were confirmed by Polymerase Chan Reaction (PCR), using previously published protocols. (1) Isolates were stored at -80°C on skimmed milk, tryptone, glucose and glycerine (STGG).

**Identification of *Salmonella* Isangi in neonatal unit samples**

Colonies with an unusual morphology were observed on ESBL chromogenic agar were initially presumed to be *Acinetobacter* spp. These colonies were flat, smooth, and colourless, in contrast to the typical appearance of *Acinetobacter* spp., which generally form smooth, raised, whitish to colourless colonies on this medium. These atypical colonies were provisionally recorded and stored as presumptive *Acinetobacter* spp. for further investigation. Following notification of a suspected *Salmonella* outbreak in the neonatal ward, a known *Salmonella Typhi* isolate was inoculated onto ESBL chromogenic agar for comparison. The colony morphology of this *S. Typhi* isolate was consistent with that of the previously stored atypical colonies. From this point onward, isolates displaying this colony morphology were recorded and stored as presumptive *Salmonella* spp. A total of 53 isolates with this morphology were archived. Subsequent testing using the Salmonella specific *ttr* PCR assay confirmed *Salmonella* spp. in 49 of these 53 isolates.

**DNA Extraction**

Isolates for DNA extraction were defrosted and plated on ESBL chromogenic agar at 37 ℃ for 18 – 24 hours. A single colony was then inoculated into 5ml of buffered peptone water at 37 ℃ for 18 – 24 in 15ml Falcon tubes. The bacterial cells were then concentrated by centrifugation at 10,000g, DNA extracted using the QIAsymphony SP (QIAGEN GmbH, Germany) automated system and the QIAsymphony DSP Virus/Pathogen Mini Kit with onboard lysis.

**Biofilm Formation**

Non-treated polystyrene 96-well plates (Corning #3370) were used for biofilm formation with or without 5 mg/mL cholesterol (Sigma #C8667) coating that had been dissolved in equal parts ethanol and isopropanol (Sigma) and then evaporated in a biosafety cabinet. *Salmonella* cultures were grown in Tryptic Soy Broth (TSB) at 37°C overnight (O/N). These cultures were normalized to an OD_600_ = 0.8 (OD_600_, optical density at 600 nm) in 1:20 TSB then further diluted 1:10 into 1:20 TSB with or without 1% human bile.(3) From this final dilution, 100 µL was dispensed in quadruplicate in 96-well plates with or without cholesterol coating. The cultured plates were incubated for 24 hours at 25°C on a GyroMini nutating mixer (LabNet International, Inc.) at 24 rpm.

After 24, biofilms attached to the 96-well plates were washed twice by submerging in buckets filled with double-distilled water (ddH_2_O), tapped excess water out onto paper towels, heat-fixed for 1 h at 60°C, and stained with 100 µL of 0.33% crystal violet per well for 5 min. After two subsequent washes in ddH_2_O, the dye was released using 100 µL of 33% acetic acid per well. To determine the intensity of staining, which correlates to the amount of biofilm present, the OD_570_ was measured in a SpectraMax spectrophotometer with SoftMax Pro software (Molecular Devices). Experiments were performed in triplicate.

**Congo red RDAR morphology**

YESCA agar plates (1 g/L yeast extract [Fisher BioReagents], 10 g/L Casamino Acids [Difco Laboratories], 15g/L agar [Fisher BioReagents]) supplemented with 40 μg/mL Congo red (Fisher Chemical) and 20 μg/mL Coomassie brilliant blue (Fisher BioReagents) were used to determine RDAR morphology. Cultures grown O/N in TSB were normalized to OD_600_ = 0.8 in water, of which 3 μL was spotted onto these agar plates, incubated at 25°C or 37°C for 4 days, and then images were captured. (4)

**Calcofluor white staining of cellulose**

Luria-Bertani (LB) no salt agar plates (10 g/L bacto tryptone [Gibco], 5 g/L yeast extract [Fisher BioReagents], 15g/L agar [Fisher BioReagents]) supplemented with 20 μg/mL Calcofluor (Fluorescent brightener 28, MP Biomedicals #158067) were used to determine cellulose levels. Plates were protected from light, as calcofluor white is light sensitive, during storage and culture growth. Cultures grown overnight in TSB were normalized to OD_600_ = 0.8, of which 3 μL was spotted onto these agar plates, wrapped in foil, incubated at 25°C or 37°C for 4 days in the dark, and then imaged with the agar facing down over a UV light source. (4) The software package Fiji within Image J was used to quantify the relative intensity levels.

**Confocal microscopy of biofilms**

*Salmonella* biofilms were grown as described above with the following modifications: O/N cultures were diluted to OD_600_ = 0.8 in 1:20 TSB and then further diluted 1:10 in 1:20 TSB. From this final dilution, 200 µL was dispensed in triplicate in 8 well chambered cover glass (Cellvis, C8-1.5H-N) without cholesterol coating. Biofilms were then grown statically at 25°C.

After 24 hours, the medium was removed from the chambered cover glass and the biofilms were carefully washed with 200 μL 1× phosphate-buffered saline (PBS); all subsequent steps should be performed slowly, to not disturb the biofilm. *Salmonella* cells and cellulose were labeled with Styo9 (5 μM; Molecular Probes) and calcofluor white (CW, 3 mg/mL; Sigma-Aldrich), respectively, in 200 μL 5% bovine serum albumin (BSA) blocking buffer at room temperature for 30 min. After incubation, the stain was removed and discarded.

To detect curli amyloid fibers, a major component of the extracellular matrix (5, 6, 7, 8, 9, 10) biofilms were then incubated with 200 μL primary rabbit α-curli antisera (1:500 in 5% BSA; courtesy of Çagla Tükel, Temple University (11) at room temperature for 30 min. The primary antibody solution was removed and discarded, washed once with 200 μL 1× PBS, and then incubated with 200 μL secondary Alexa Fluor 633 goat anti-rabbit IgG (1:1,000 in 5% BSA; Invitrogen) at room temperature for 30 min. The secondary antibody was removed and discarded, and the biofilms were fixed with 200 μL 4% paraformaldehyde (PFA, Affymetrix) at room temperature for 30 min prior to confocal imaging. All steps were carried out with as little light exposure as possible and covered with foil to prevent photobleaching.

Using a Zeiss LSM 800 confocal laser scanning microscope with a Plan-Apochromat 63x/1.4 Oil DIC M27 objective, stained biofilms were visualized, and three-dimensional biofilm images were acquired by capturing 2 random Z-stacks per well, 3 wells per strain. The signal for each fluorophore was recorded separately: Syto9-labeled *Salmonella* were visualized at an excitation wavelength of 483 nm and an emission wavelength of 500 nm, calcofluor white-bound cellulose was visualized at an excitation of 254 nm and an emission of 432 nm, and Alexa Fluor 633-labeled curli fimbriae were visualized at an excitation of 631 nm and an emission of 647 nm. The Z-stacks were then analyzed using the software package Comstat2 within Image J to calculate biomass, average thickness, and maximum thickness (12).

**Mouse Infection**

To test virulence, we infected eight week old (16 female) BALB/c mice (Jackson Laboratory, Bar Harbor, ME) intragastrically (IG) with 1x10^5^ CFU/mL in PBS or 100 µL PBS (4 each) and monitored for 14 days, euthanizing when moribund.

**MIC/MBC**

To determine the minimum inhibitory concentration (MIC) of common disinfectants utilized in a hospital setting against *Salmonella*, a broth microdilution method was used, performed in non-treated polystyrene 96-well plates (Corning). Cultures were grown O/N in Cation Adjusted Mueller Hinton Broth (CAMHB). The following day, 100 µL was subcultured into 4.9 mL fresh CAMHB for 4-6 hours rolling at 37°C, and subsequently adjusted to an OD_600_ = 0.0003 (~ 5 x 10^5^ CFU/mL) in 2 mL CAMHB for each strain and disinfectant to be tested. From the 2 mL, 500 µL was transferred to a new tube, to which the maximum concentration of disinfectant to be tested was added, just before being added to the plate and serially diluted: 1% bleach (Chlorox concentrated germicidal bleach, Grainger #41H893), 240 µg/mL chlorine (SpaGuard chlorinating concentrate, Amazon #B007ZU460S), 0.125% chlorhexidine gluconate (Hibiclens 4.0% w/v solution, Amazon #B00EV1D79A). Column 12 was filled with 100 µL CAMHB as a negative control and columns 2-11 with 100 µL of the remaining 1.5 mL bacterial culture. Immediately upon addition of disinfectant to the 500 µL aliquot, column 1 was filled with 200 µL and immediately 100 µL was removed and mixed 10 times into column 2, and continued to mix quickly but carefully through column 10, removing a final 100 µL from column 10 after being mixed which results in equal volumes in all wells (leaving column 11 as the positive control with bacteria only). The 96-well plates were wrapped in parafilm, placed in a lidded plastic container with damp paper towels, and incubated for 16-18 hours at 37°C. To determine lowest concentration in which less than 10% growth occurred, the OD_600_ was measured in a SpectraMax spectrophotometer with SoftMax Pro software (Molecular Devices). The percentage of inhibition was determined by comparing the positive control (column 11) after subtraction of the negative control (column 12).

To determine the minimum bactericidal concentration (MBC), before the MIC was recorded, the 96-well plates were gently mixed on a basic vortex mixer fitted with a microplate tray attachment (500 rpm for 1 minute, Thermo Scientific), then 3 µL from each well was spotted onto LB agar, allowed to dry, and incubated for 24 hours at 37°C. The dilution with the lowest concentration that produced no growth was recorded as the MBC.

**Microbiologic methods for recovering salmonellae in South Africa**

**Surveillance for clinical isolates of *Salmonella* in South Africa**

The National Institute for Communicable Diseases (NICD) is a national public health institute for South Africa, providing disease surveillance, specialised diagnostic services, outbreak response, public health research and capacity building to support the government’s response to communicable disease threats. The Centre for Enteric Diseases (CED), NICD plays a part in national laboratory-based surveillance for human isolates of *Salmonella*. As part of this “GERMS-SA Laboratory Surveillance Network”, the CED receives isolates from ~200 public and private clinical microbiology laboratories throughout the country.

**Receipt of bacterial cultures and phenotypic characterization**

Following receipt of presumptive *Salmonella* isolates on Dorset-Egg transport media [Diagnostic Media Products (DMP), National Health Laboratory Service, Johannesburg, South Africa], isolates are sub-cultured onto 5% Blood Agar (DMP) to check for viability and purity, following which the isolates are processed to extract genomic DNA for WGS analysis. If there is suspicion that a culture is not a *Salmonella*, then that culture will be further investigated using standard phenotypic microbiological identification and serotyping methodologies, including VITEK-2 identification (bioMérieux, Marcy-l'Étoile, France) and serotyping completed as per the White-Kauffmann-Le Minor Scheme. When required, antimicrobial (ampicillin, ciprofloxacin, ceftriaxone, azithromycin) susceptibility testing was achieved via Etest methodology (bioMérieux).

**Genomic DNA extraction and WGS of bacteria**

Genomic DNA was extracted from bacteria using either the Qiagen QIAamp DNA Mini Kit (QIAGEN, Hilden, Germany) or the Invitrogen PureLink Microbiome DNA Purification Kit (Invitrogen, Waltham, Massachusetts, USA). WGS was performed by the NICD Sequencing Core Facility, using Illumina NextSeq technology (Illumina, San Diego, CA, USA). DNA libraries were prepared using the Illumina DNA Prep Kit. Sequencing included paired-end sequencing runs, including ~80 times coverage.

**Environmental isolate processing**

Environmental isolates were processed as per Rigby et al., 2025^2^.

Supplementary Table 1 . The date and source of 37 isolates collected during the ChatinkhaRes research study at Queen Elizabeth Central Hospital

| **Isolate ID** | **Ward** | **Collection date** | **Site of origin** |
| --- | --- | --- | --- |
| CIS153C1 | Chatinkha nursery | 11/02/2020 | Chitenje |
| CIS15HB1 | Chatinkha nursery | 04/03/2020 | Cot |
| CIS15MR1 | Chatinkha nursery | 16/03/2020 | Child Rectal |
| CIT159B1 | Chatinkha nursery | 28/02/2020 | Cot |
| CIT15EB1 | Chatinkha nursery | 09/03/2020 | Cot |
| CIT15FB1 | Chatinkha nursery | 13/03/2020 | Cot |
| CIT15FH1 | Chatinkha nursery | 13/03/2020 | Mother Hand |
| CIT15GH1 | Chatinkha nursery | 27/03/2020 | Mother Hand |
| CIT15GS1 | Chatinkha nursery | 19/03/2020 | Child Stool |
| CIU13XS1 | Chatinkha nursery | 17/02/2020 | Child Stool |
| CIU145B1 | Chatinkha nursery | 04/03/2020 | Cot |
| CIU149H1 | Chatinkha nursery | 13/03/2020 | Mother Hand |
| CIV13PE7 | Chatinkha nursery | 17/02/2020 | Surfaces surgical bay |
| CIV13RE2 | Chatinkha nursery | 24/02/2020 | Side room 2 |
| CIV13UE4 | Chatinkha nursery | 02/03/2020 | Metal sink |
| CIV13VE5 | Chatinkha nursery | 02/03/2020 | Cots surgical bay |
| CIV13XE2 | Chatinkha nursery | 09/03/2020 | Side room 1 |
| CIV13XE5 | Chatinkha nursery | 09/03/2020 | Handwash sink |
| CIV13XE8 | Chatinkha nursery | 09/03/2020 | CPAP |
| CIV13XU1 | Chatinkha nursery | 09/03/2020 | Nurses station |
| CIV13YE6 | Chatinkha nursery | 09/03/2020 | Sats probes |
| CIV13YE8 | Chatinkha nursery | 09/03/2020 | Floor |
| CIV140E1 | Chatinkha nursery | 16/03/2020 | Side room 2 |
| CIV140E4 | Chatinkha nursery | 16/03/2020 | Patient chairs |
| CIV140E6 | Chatinkha nursery | 16/03/2020 | CPAP |
| CIV140E7 | Chatinkha nursery | 16/03/2020 | Solid surfaces |
| CIV140U1 | Chatinkha nursery | 16/03/2020 | Side room 1 |
| CIV141E6 | Chatinkha nursery | 16/03/2020 | Sats probes |
| CIV142E6 | Chatinkha nursery | 16/03/2020 | Oxygen equipment |
| CIV143E2 | Chatinkha nursery | 23/03/2020 | Side room 2 |
| CIV143E4 | Chatinkha nursery | 23/03/2020 | Handwash sink |
| CIV143E9 | Chatinkha nursery | 23/03/2020 | Nurses station |
| CIV143U1 | Chatinkha nursery | 23/03/2020 | Supply trolley |
| CIV144E6 | Chatinkha nursery | 23/03/2020 | Scales |
| CIV144U1 | Chatinkha nursery | 23/03/2020 | Patient sinks |
| CIV146E8 | Chatinkha nursery | 30/03/2020 | Solid surfaces |
| CIV147E3 | Chatinkha nursery | 30/03/2020 | Surfaces surgical bay |

Supplementary Table 2. Minimum Inhibitory Concentrations (MIC) and Minimum Bactericidal Concentrations (MBC) of Bleach, Chlorine, and Chlorhexidine for Salmonella Isolates. Bleach and chlorine were bactericidal for all isolates (MIC = MBC). Salmonella Isangi (CAAXQV, CAAYM5, CIV143E9) exhibited lower MICs to chlorhexidine compared to Salmonella Typhimurium ST19/14028. However, ST19 4/74 and one Isangi isolate required slightly higher chlorhexidine concentrations for bacterial killing (MBC > MIC).

| **Isolate** | **MIC Bleach (%)** | **MIC Chlorine (µg/mL)** | **MIC Chlorhexidine (%)** | **MBC Bleach (%)** | **MBC Chlorine (µg/mL)** | **MBC Chlorhexidine (%)** |
| --- | --- | --- | --- | --- | --- | --- |
| **14028** | 0.03125 | 120 | 0.001953125 | 0.03125 | 120 | 0.001953125 |
| **ST19** | 0.03125 | 120 | 0.001953125 | 0.03125 | 120 | 0.00390625 |
| **CAAXQV** | 0.03125 | 120 | 0.000946563 | 0.03125 | 120 | 0.001953125 |
| **CAAYM5** | 0.03125 | 120 | 0.000946563 | 0.03125 | 120 | 0.00390625 |
| **CIV143E9** | 0.03125 | 120 | 0.000946563 | 0.03125 | 120 | 0.001953125 |

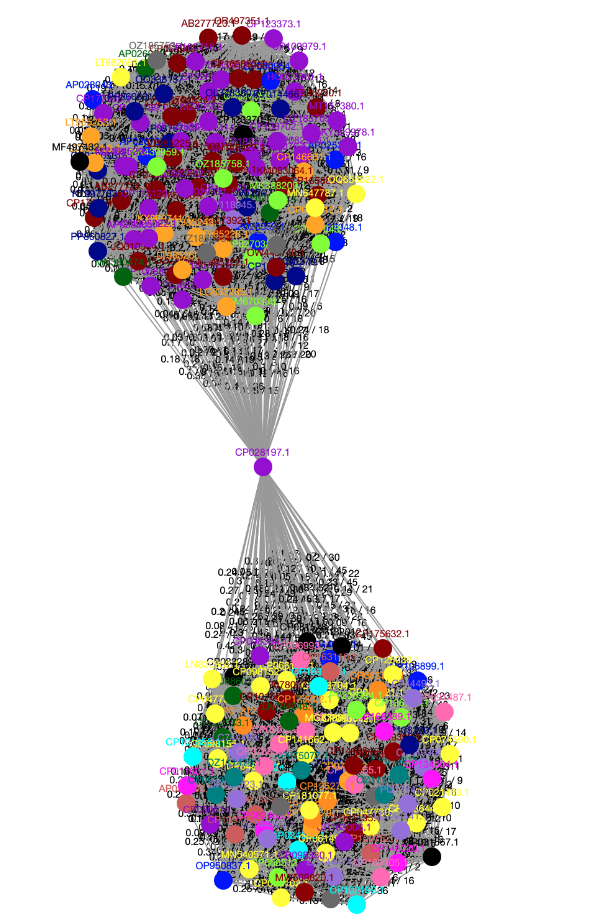

Supplementary Figure 1. Plasmid similarity network from pling, based on DCJ-Indel distance. Each node is a single plasmid, and the edges are DCJ-Indel distance. The two densely connected clusters are IncC plasmids and IncHI2 plasmids, the central node (CP028197.1) connects both clusters as it is a cointegrate plasmid that is closely related by DCJ-Indel distance to both plasmid backbone

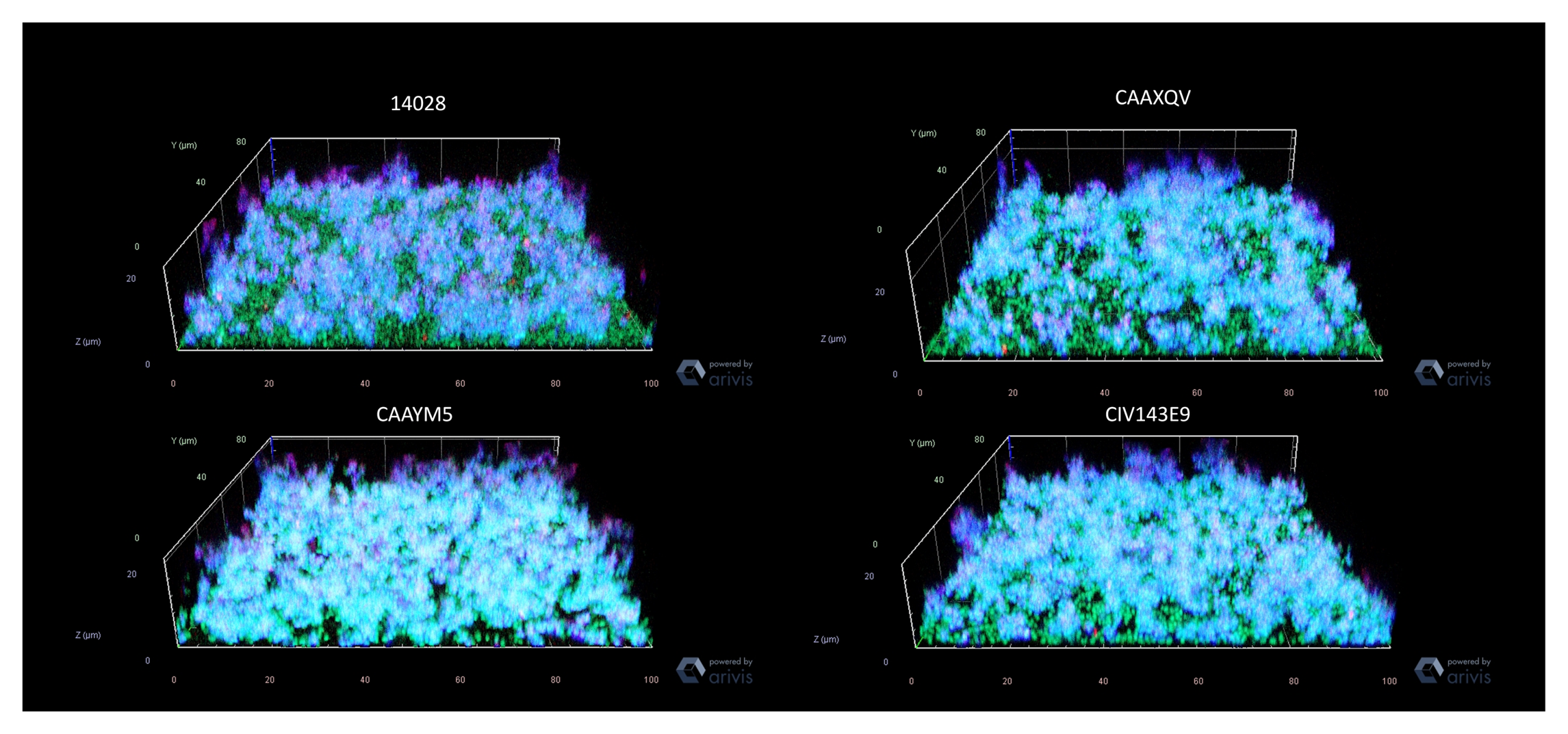

Supplementary figure 2. Differences in biofilm formation visualized by confocal microscopy. Pictured are representative Z-stack images of WT Salmonella Typhimurium ATCC 14028 or selected Salmonella Isangi isolates CAAXQV, CAAYM5, CIV143E914028 and the three Isangi strains. Isolates were grown statically in chambered coverglass slides in 200 μL 1:20 TSB statically at 25°C for 24 hours. Planktonic cells were washed away then Syto-9 (viable cells-green), calcofluor white (cellulose-blue), and rabbit α-amyloid monoclonal antibody (Mab)(curli amyloid fibers-red) were used to visualize the biofilm prior to fixation with 4% paraformaldehyde (PFA). Representative Z-stacks (2 stacks/well, 3 wells/strain) were captured with the 63x/1.4 Oil objective on a Zeiss LSM 800 laser scanning confocal microscope. Despite the variability in cellulose expressed on CW plates, confocal imaging of 24 hour biofilms showed no discernable difference.

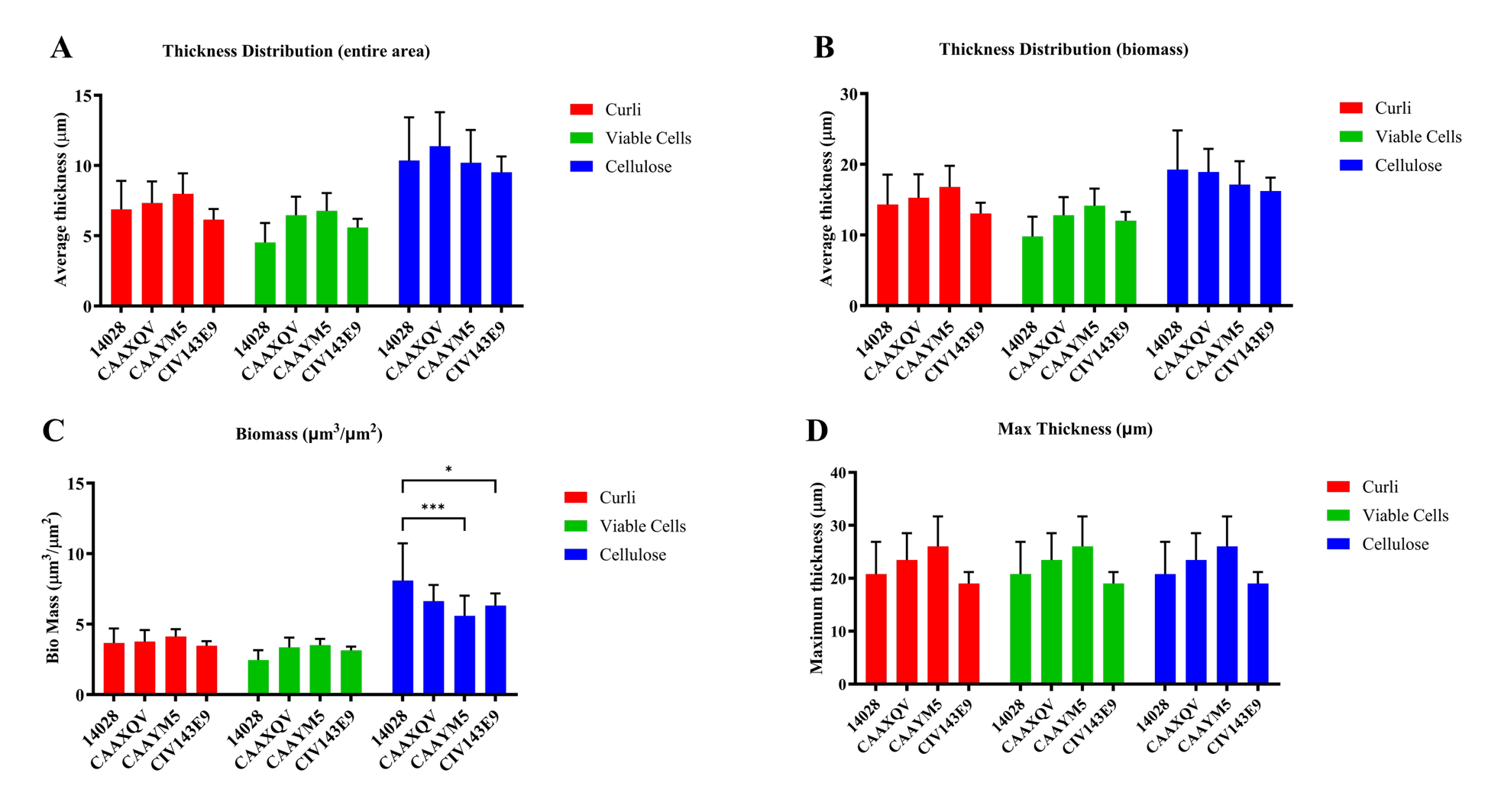

Supplementary figure 3. Quantification of biofilm formation visualized by confocal microscopy. Wild type *Salmonella* Typhimurium ATCC 14028 or selected Salmonella Isangi isolates CAAXQV, CAAYM5, CIV143E9 were grown statically in chambered coverglass slides in 200 μL 1:20 TSB statically at 25°C for 24 hours. Planktonic cells were washed away then Syto-9 (viable cells-green), calcofluor white (cellulose-blue), and rabbit α-amyloid monoclonal antibody (Mab)(curli-red) were used to visualize the biofilm prior to fixation with 4% paraformaldehyde (PFA). Representative Z-stacks (2 stacks/well, 3 wells/strain) were captured with the 63x/1.4 Oil objective on a Zeiss LSM 800 laser scanning confocal microscope. Comstat2 software was used to calculate the thickness distribution (A and B), biomass (C), and maximum thickness (D) of biofilm Z-stacks for each individual component (cells, cellulose, and curli). Data are presented as the mean ± SD. *, P < 0.05; ***, P < 0.001.

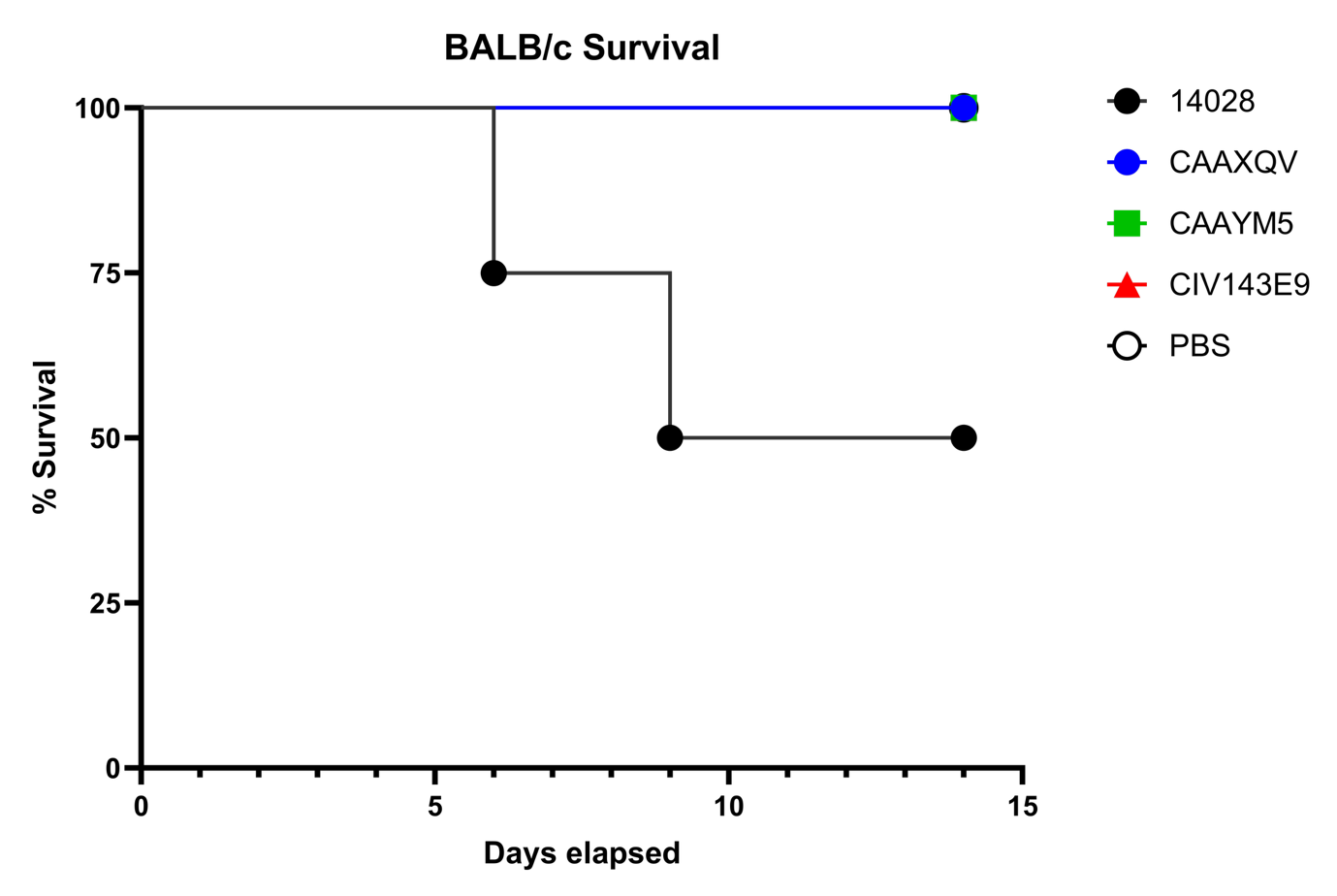

Supplementary Figure 4. Survival of BALB/c mice following intragastric infection with Salmonella isolates. Eight-week-old female BALB/c mice were infected intragastrically (1×10⁵ CFU/mL) with Salmonella Typhimurium ATCC 14028, selected Salmonella Isangi isolates (CAAXQV, CAAYM5, CIV143E9), or PBS. S. Isangi infection did not result in mortality, whereas S. Typhimurium 14028 caused 50% mortality by day 14.
